# Vital Insights: How Radar-Based Monitoring Can Enhance Symptom Management in Palliative Care

**DOI:** 10.1101/2025.04.02.25325074

**Authors:** Thanh Trúc Trần, Julia Yip, Marie Oesten, Stefan G. Griesshammer, Nils C. Albrecht, Maria Heckel, Bjoern M. Eskofier, Alexander Koelpin, Christoph Ostgathe, Tobias Steigleder

**Affiliations:** Machine Learning and Data Analytics Lab, Friedrich-Alexander-Universität Erlangen-Nürnberg (FAU), Erlangen, Germany; Department of Palliative Medicine, Universitätsklinikum Erlangen, Friedrich-Alexander-Universität Erlangen-Nürnberg (FAU), Erlangen, Germany; Institute for High Frequency Technology, Hamburg University of Technology (TUHH), Hamburg, Germany

**Keywords:** Palliative Care, Symptom Management, Radar Technology, Vital Signs

## Abstract

**Context:** Palliative care prioritizes patients’ quality of life with symptom management, personal care, and minimal technology use to respect privacy. However, assessing symptoms, particularly for patients near the end of life who often are unable to communicate, mostly depends on observation by healthcare providers, which may lack completeness and objectivity.

**Objectives:** Non-invasive radar-based monitoring of heart rate (HR) and respiration rate (RR) offers potential for detecting symptoms without compromising palliative care principles.

**Methods:** We used a multi-radar system placed under patients’ mattresses to continuously monitor HRs and RRs in a palliative care ward by tracking body surface movement. Data on symptom burden, including timing and severity, were collected via patient self-reports (MIDOS^2^) and staff observations (HOPE). The relationship between radar-derived vital signs and distress-related symptom burden changes over hours, as well as medication effects following their peak impact, was analyzed using various statistical methods.

**Results:** Our findings show that HR and RR increase over the course of hours during symptom exacerbation (HR: +2.8 bpm, RR: +0.7 brpm) and decrease with amelioration (HR: –1.8 bpm, RR: –0.2 brpm), with a significant relationship observed between HR and symptom exacerbation (p=0.002). Additionally, HR trends suggested variability potentially related to medication effects, warranting further study.

**Conclusion:** Radar-based monitoring holds promise for burden-free vital sign monitoring in palliative care and assisting healthcare professionals in symptom detection. However, physical activity and environmental factors may affect measurements and medication effects require further investigation. Additional research is needed to refine this technology for clinical use.

## 1. Introduction

Palliative care (PC) focuses on improving the quality of life for patients facing life-limiting or life-threatening illnesses in a holistic manner by symptom amelioration, promoting social inclusion, communication, and personal care[1]. Realizing these principles requires accurate detection of detrimental factors, such as symptom burden, and indicators crucial for tailored therapy planning, like health status trajectories and lifetime prognosis[2]. Currently, symptom assessment primarily relies on self-reports by patients. However, this method has limitations: while some symptoms like pain are regularly reported, others – such as dyspnea, loss of appetite, or sadness/depression – are mentioned less frequently, likely due to social expectancy and biases[3]. Moreover, over half of the patients in PC wards, and nearly all patients in their final hours or days of life, are unable to communicate their symptoms[4].

The practice of assessment based on observations by health care professionals (HCP) to predict symptom burden and estimate health trajectories, introduces a substantial degree of bias[5, 6]. Such interpretations are inherently subjective and transient, and thus fail to provide a comprehensive picture. Furthermore, the presence of HCP may influence the behavior of the patient, making the observations less reliable. Therefore, it is necessary to consider more objective assessment methods. In other medical fields, such as intensive care, continuous monitoring devices provide valuable data on patients’ health status and distress indicators, such as rising blood pressure, heart rate (HR) and respiratory rate (RR). However, in PC, such technological interventions are often inappropriate as they contravene the fundamental principles of social inclusion, communication, and personal care. Avoiding these established medical devices presents a major challenge: while continuous, objective data is essential for optimal patient care, the methods of obtaining this data must not compromise the therapeutic setting.

Innovative, touchless technologies for assessing vital parameters may present a viable alternative, as they are designed to preserve patient autonomy and are less likely to disrupt relationship-centered care or the provision of psychosocial support. For example, electro-optical and video-based approaches[7] capture subtle physiological changes, such as skin color variations indicative of HR and chest movements for respiration. However, these methods give rise to concerns regarding privacy, as continuous video monitoring may be perceived as intrusive. Furthermore, they require a free line-of-sight and consistent lighting conditions. Infrared technology [8, 9], which detects heat patterns to monitor HR and breathing, shares similar limitations regarding line-of-sight and environmental lighting, potentially introducing artifacts. Another concept of long-term monitoring is ballistocardiography[10] which detects mechanical vibrations resulting from cardiac and respiratory activities through sensors embedded in surfaces such as mattresses. Although this approach is less intrusive, it requires direct contact with the surface and is highly sensitive to movement, which can compromise accuracy, introduce artifacts or trigger false alarms.

Despite these advancements, there remains a critical need for a touchless, burden-free monitoring solution that guarantees privacy and operates independently of line-of-sight and lighting conditions, while ensuring reliable vital signs monitoring (VSM) for both inpatient and outpatient care. Radar technology is emerging as a promising solution[11]. In contrast to the aforementioned methods, radar is not constrained by visual obstruction and capable of penetrating various materials such as wood, clothing, and mattresses. This enables the discreet installation of radar systems beneath mattresses or behind wooden panels while still ensuring the continuous measurement of movements caused by the heartbeat or breathing[12]. Considerable progress has been made in this area, indicating a potential pathway to integrating continuous, objective health monitoring in PC without compromising its core therapeutic principles[13].

While previous research mostly focuses on obtaining vital parameters from radar data, this study is the first to investigate how symptom burden of PC patients in a real-world setting is reflected in radar-based assessments of HR and RR. By employing radar technology, our objective is to develop a continuous, unobtrusive VSM approach that has the potential to transform symptom management by assisting HCPs in tailoring therapeutic approaches to individual patient needs.

The present study specifically investigated the ability of radar-based HR and RR assessment in reflecting symptom trajectories and responses to medication in PC patients, with particular focus on distress-related symptoms. In the context of comparable clinical settings and experience with VSM, an increase in HR/RR is typically observed when patients experience distressing symptoms via a sympathetic activation, followed by a decrease when symptoms are ameliorated via reduced sympathetic activity and increased parasympathetic activity[14, 15]. The study was conducted under real-life conditions on a PC ward with the objective of evaluating the potential of radar technology as a non-intrusive, continuous monitoring tool in this sensitive patient population for future clinical application. The findings suggest that radar-based monitoring provides valuable insights into patient well-being, offering a new and promising avenue for enhancing PC through improved symptom management and medication effectiveness assessment.

## 2. Methods

### 2.1. Study Design

This prospective observational study was conducted in the PC unit of the Universitätsklinikum Erlangen, Germany, with the approval of the local ethics committee (479_20 B), to ensure compliance with ethical standards in PC and research. Participant inclusion criteria were age over 18 years and informed consent was obtained from the patient themselves or their legal representative before starting the trial. The study is registered in the Bavarian Cancer Research Center under the acronym GUARDIAN [16].

Twelve beds were equipped with a multi-radar system as described below. The duration of radar data collection varied according to the length of patient stay. After extracting HRs and RRs from the radar signal as explained below, we used a long-term and a short-term approach to assess the correlation between vital signs and symptom burden. Long-term analysis tracked symptom trajectories and radar-based vital parameters over the course of several hours or days. Short-term analysis focused on the effect of on-demand medication by measuring vital signs before administration and after the medication’s peak effectiveness was reached.

Symptom data were collected using the “Minimal Documentation System” (MIDOS^2^)[17] for PC patient self-reports after peak effectiveness of on-demand medication, and the “Hospice and Palliative Evaluation” (HOPE)[18] questionnaire for staff observations which were conducted typically three times a day. In both MIDOS-2 and HOPE, the severity of each symptom is rated on a four-step scale, with 0 indicating no symptom occurrence and 3 representing strong symptom severity. For long-term analysis, symptom severity changes were tracked across multiple consecutive HCP observations, with symptom improvement defined as a decrease of –2 or –3, and symptom exacerbation as an increase of +2 or +3. Intermediate changes or fluctuations were excluded from the analysis. For short-term analysis, medication was classified as effective if the patient reported symptom severity as 0 or 1 after the period of peak effectiveness, whereas a severity of 2 or 3 was classified as ineffective.

In consideration of the distinctive context of a PC ward, where patients are approaching the end of life, ethical considerations and the principle of beneficence were prioritized to maintain a normal and minimally intrusive environment for patients and their families. Consequently, no additional validation technologies, such as ECG or other monitoring probes, were employed. Additionally, ground truth data from clinical health records (CHR) was collected retrospectively. While this approach yielded valuable insights, limitations such as potentially imprecise timestamp accuracy must be acknowledged. Nonetheless, all information remains valid within clinically relevant parameters.

### 2.2. Multi-Radar System

A radar-based remote sensing system for heartbeat and respiration detection was employed, allowing under-mattress placement at the level of the patients’ chests[19, 20]. To ensure continuous VSM across the entire width of the bed, the system incorporated four continuous wave (CW) radars with even spacing, each operating on distinct frequencies within the 24GHz band to prevent crosstalk. The spacing aligned with the holes in the hospital bed frame (Figure 1b). Thus, an 18 cm foam mattress, as well as bedding and clothing are located between the radar antennae and the surface of the patient’s body. The CW signals can penetrate these materials unchanged due to minimal variations in permittivity. The half-power beamwidth of the bistatic antenna setup beamwidth was 34° and 49°, respectively (Figure 1a). Partial intersection of the radar beams across the entire mattress surface ensured maximal signal coverage and consistent data acquisition for all patient positions.

**Figure 1:**
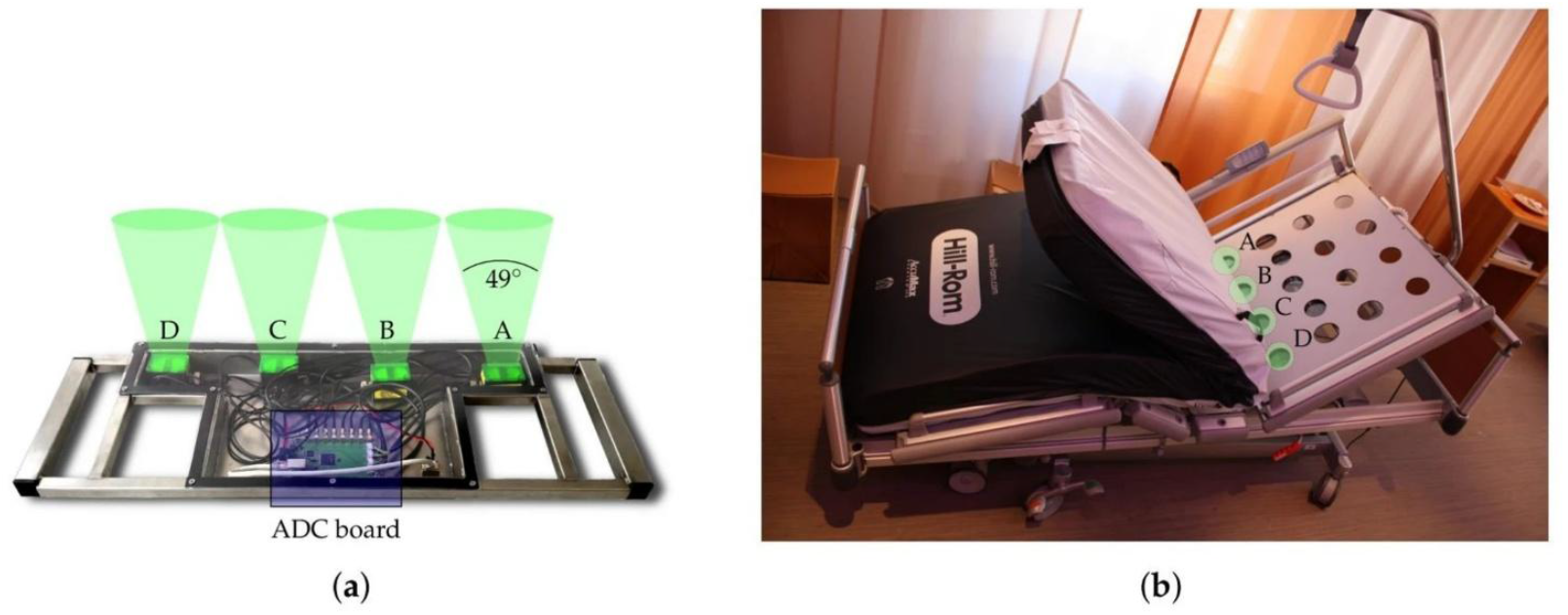
(a) Radar system with four evenly spaced CW modules and ADC (Analog to Digital Converters) board. (b) Hospital bed with holes aligning with radar modules. (Source: Schellenberger et al. [19])

### 2.3. Vital Sign Extraction

An ellipse reconstruction technique as described in [21] was employed to mitigate offset voltages in the in-phase (I) and quadrature (Q) radar signals. Using the phase shift (φ) between the transmitted and received signals, the relative displacement (x(t)) was calculated, allowing for the retrieval of heartbeat and respiration information.

HRs are typically extracted by analyzing the radar signal’s frequency spectrum containing the pulse wave[22, 23]. However, isolating the heartbeat component from the overall pulse wave is challenging due to interference from respiration and body movements, especially in short observation windows[24]. Thus, we estimated HRs by detecting individual heart sounds (HS) as described in [25] instead, which offers instantaneous heartbeat detection and improved accuracy. A fourth-order bandpass Butterworth filter encompassing the HS range (16–80 Hz) was applied, followed by a Hidden Semi-Markov Model (HSMM)-based algorithm to estimate the four distinct states of the HS: first heart sound (S1), systole, second heart sound (S2), and diastole[26]. Interbeat intervals (IBIs) were extracted from the transitions between these distinct states, particularly the onsets of S1 and S2. HRs were obtained by inverting the IBI values.

Respiratory information was extracted by passing the radar signal through a fourth-order Butterworth filter (0.07–0.7 Hz) and analyzing zero crossings from positive to negative values, corresponding to chest wall movements during breathing[20]. The intervals between zero crossings were used to calculate RR.

### 2.4. Outlier Detection

While HSMMs offer a promising approach for HR estimation from radar-based HSs, their accuracy heavily relies on correctly detecting and differentiating between S1 and S2. The HSMM used in this study was trained on radar data collected in a controlled laboratory environment without motion artifacts[27]. However, in clinical settings like this one, patients were less likely to remain static, introducing noise from random body movements into the radar signal and complicating reliable detection of HS states. This can result in several seconds of incorrect HS measurements when a state is missed[28], ultimately leading to inaccurate HR estimation in this timeframe. To address this problem, we implemented a two-stage iterative outlier detection process to improve HR accuracy, striking a balance between eliminating genuine outliers while preserving valid HR data that exhibits some natural variability, particularly with increases during physical activity or distress as exemplary shown in Figure 2.

**Figure 2:**
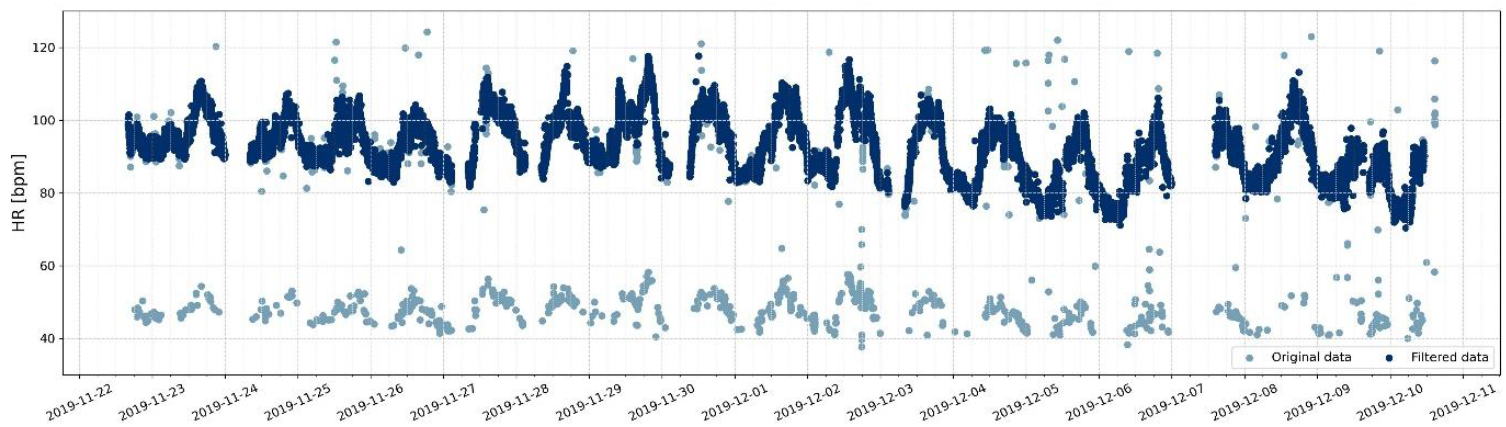
Exemplary radar-based HR trajectory before and after outlier removal.

While the specific two-stage process described here was developed independently due to the lack of an existing method suitable for this study’s needs, it draws on established outlier detection techniques that combine global and local approaches (e.g., [29, 30, 31]). First, global outliers were removed by comparing data points to the median HR over the entire period, excluding points outside a median-dependent threshold. The second stage targeted local outliers, while still accounting for potential spontaneous fluctuations in HR. A localized iterative sliding time window recalculated the median within each window, strategically reducing the window size and minimum required number of observations to capture localized outliers while preserving realistic HR fluctuations. A sliding window Z-score method further refined the data, with HR points lacking valid neighbors classified as outliers.

### 2.5. Data Analysis

The relationship between symptom changes and radar-derived vital parameters was examined through long-term and short-term analyses, both using HR/RR trajectories consisting of exactly one start and one end point. Long-term analysis focused on significant symptom changes over the course of several hours with their start and end times documented in CHR. Short-term analysis assessed the effects of demand medication, where the start time was when the medication was administered, and the end time was based on expected peak effect: 60 minutes for oral, 30 minutes for subcutaneous, and 10 minutes for intravenous administration. Figure 3 illustrates an exemplary HR short-term trajectory, showing the period from the administration of hydromorphone (subcutaneous) for dyspnea to the expected peak effect. HR/RR values were averaged within ±3 minutes around the start and end of each symptom trajectory. Analyses included chi-squared tests, pairwise t-tests, linear regression, and generalized linear mixed models (GLMMs) to account for repeated measures and uneven data distribution.

**Figure 3:**
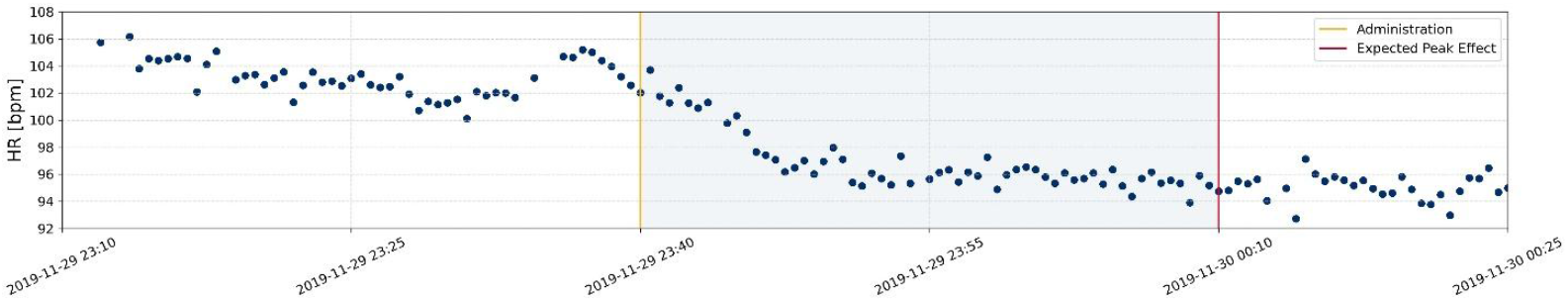
Exemplary HR short-term trajectory between administration of on-demand medication (hydro-morphone (subcutaneous) for dyspnea) and expected maximum effect.

## 3. Results

### 3.1. Dataset

Seventy-seven palliative care patients participated in our study, with 41 female and an average age of 70.73 years. HR and RR trajectories that started or ended with values outside the predefined limits (55-130 bpm for HR and 5-30 brpm for RR) were excluded from the analysis, resulting in the examination of n=230 HR (n=111 RR) long trajectories and n=282 HR (n=116 RR) short trajectories. Long HR and RR trajectories were derived from 44 and 28 patients, respectively, while 43 and 23 patients contributed to the HR and RR short trajectories, respectively.

### 3.2. Long-term Analysis

Table 1 presents the average changes in HR and RR over long trajectories, separated into two outcome groups: symptom amelioration and symptom exacerbation. Scatterplots and boxplots in Figure 4 provide additional insights into the distribution of these symptom severity changes.

**Table 1:** Long-term analysis – Average changes in HR and RR associated with symptom amelioration and exacerbation.

|  | HR<br>Sample<br>Size | HR Before<br>(bpm) | HR After<br>(bpm) | RR<br>Sample<br>Size | RR Before<br>(brpm) | RR After<br>(brpm) |
| --- | --- | --- | --- | --- | --- | --- |
| Symptom<br>Amelioration | 118 | 88.2 ± 15.7 | 86.4 ± 15.7 | 63 | 14.0 ± 4.9 | 13.8 ± 4.1 |
| Symptom<br>Exacerbation | 112 | 85.8 ± 14.6 | 88.6 ± 16.5 | 48 | 13.7 ± 4.1 | 14.4 ± 4.6 |

**Figure 4:**
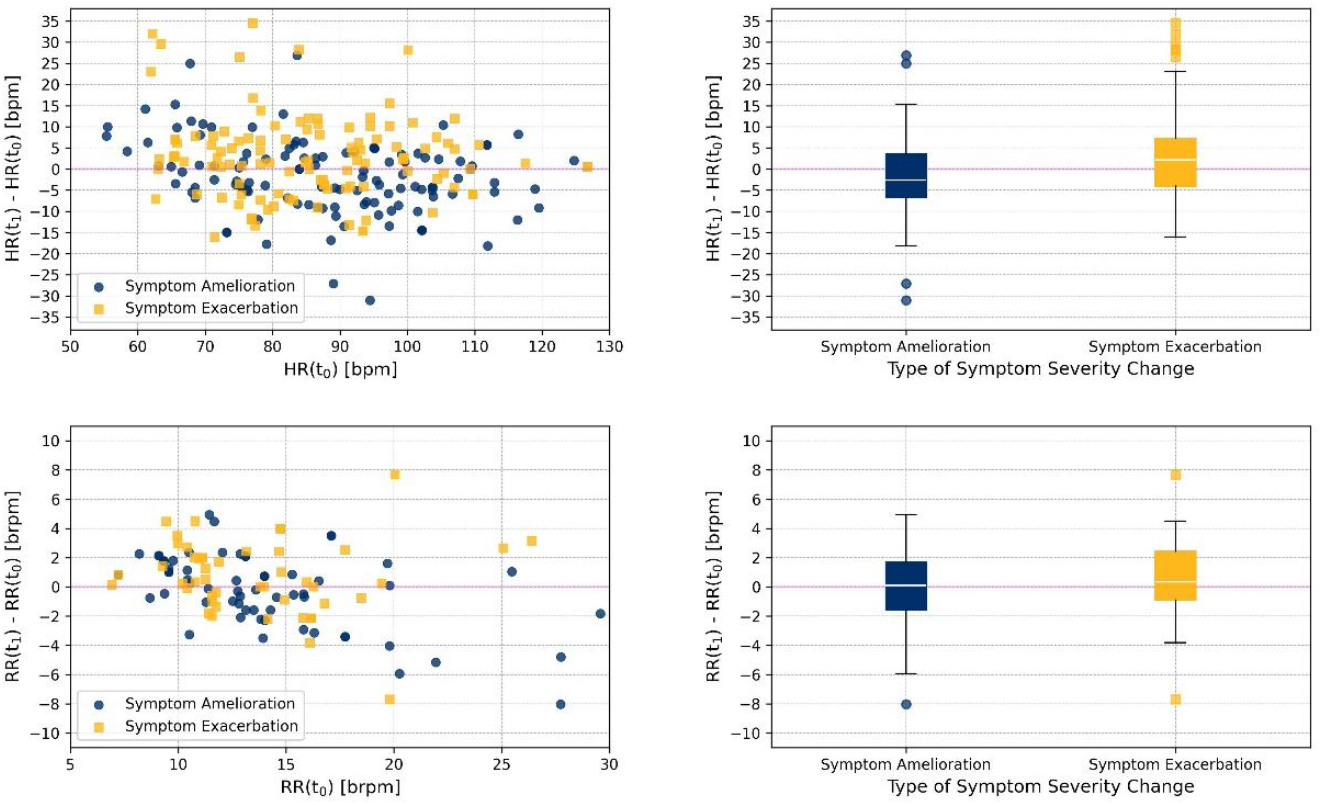
Long-term analysis – Distribution of HR and RR changes corresponding to symptom amelioration and exacerbation (t_0_: before, t_1_: after symptom severity change).

For assessing symptom burden using Pearson’s chi-square test of independence, we categorized symptom changes as amelioration or exacerbation, and classified HR/RR changes as increases or decreases. Test results indicated a significant general association between symptom severity and HR (χ^2^(1, n=230) = 8.66, p=0.003), but not with RR (χ^2^(1, n=111) = 1.59, p=0.208). Pairwise t-tests were conducted to compare the mean vital parameter changes across the two outcome groups (Table 2). Significant results were observed only in cases of symptom exacerbation (HR: p=0.002, RR: p=0.044).

**Table 2:** Long-term analysis – Pairwise t-test results for HR and RR changes by symptom amelioration and exacerbation.

|  | HR<br>Sample<br>Size | HR<br>t-value | HR<br>p-value | RR<br>Sample<br>Size | RR<br>t-value | RR<br>p-value |
| --- | --- | --- | --- | --- | --- | --- |
| Symptom<br>Amelioration | 118 | 2.196 | 0.985 | 63 | 0.731 | 0.766 |
| Symptom<br>Exacerbation | 112 | -2.975 | 0.002 | 48 | -1.746 | 0.044* |

Additionally, logistic regression was used to examine the relationship between symptom and vital parameter changes (Table 3), with results indicating a significant relationship with HR (p=0.001), while the connection with RR approached significance (p=0.078). To account for individual differences between patients andthe effect of time, GLMMswerealso performed. However, adjusting for individual patient effects did not alter the results, as the estimates and p-values for both HR and RR remained consistent with those obtained from the logistic regression. Although time differences between start and end points of the long trajectories varied considerably (HR: 13.9 ± 11.4 h; RR: 13.1 ± 12.6 h), accounting for this time dependency had minimal impact on the regression outcomes.

**Table 3:** Long-term analysis – Logistic regression and GLMM results for HR and RR changes related to symptom amelioration and exacerbation.

| Variable | Model Type | Intercept<br>Source | Intercept<br>Estimate | Intercept<br>p-value | Variable<br>Estimate | Variable<br>p-value |
| --- | --- | --- | --- | --- | --- | --- |
| HR<br>change | Logistic<br>Regression | Constant | -0.074 | 0.587 | 0.054 | 0.001*** |
| HR<br>change | GLMM | Time<br>difference | -0.086 | 0.575 | 0.061 | 0.001*** |
| RR<br>change | Logistic<br>Regression | Constant | -0.303 | 0.122 | 0.142 | 0.078 |
| RR<br>change | GLMM | Time<br>difference | -0.370 | 0.176 | 0.210 | 0.086 |

### 3.3. Short-Term Analysis

Table 4 shows radar-based HR/RR changes following demand medication to relieve distress-related symptoms. Symptom improvement was observed for over 90% in both HR and RR trajectories, leading to a strong class imbalance, as also illustrated in the scatterplots in Figure 5. Given this class imbalance, we determined that logistic regression was not suitable for our analysis.

**Table 4:** Short-term analysis – Average HR and RR changes depending on medication response.

|  | HR<br>Sample<br>Size | HR<br>Before<br>(bpm) | HR After<br>(bpm) | RR<br>Sample<br>Size | RR<br>Before<br>(brpm) | RR After<br>(brpm) |
| --- | --- | --- | --- | --- | --- | --- |
| Effective<br>Medication | 256 | 94.0 ± 15.8 | 93.2 ± 15.0 | 113 | 14.9 ± 4.8 | 14.2 ± 4.4 |
| Ineffective<br>Medication | 26 | 96.9 ± 14.8 | 98.9 ± 16.8 | 3 | 11.9 ± 0.6 | 12.9 ± 0.5 |

**Figure 5:**
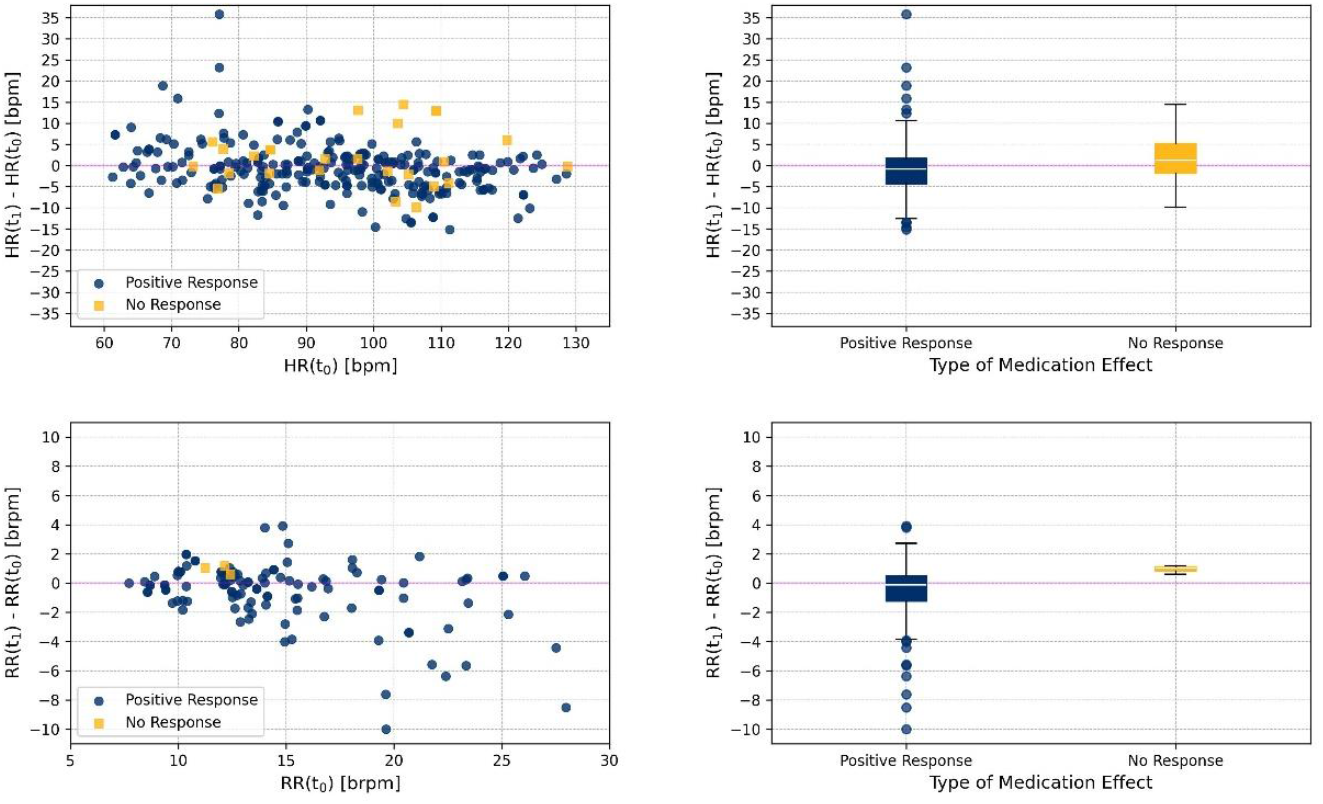
Short-term analysis – Distribution of HR and RR changes depending on response to medical intervention (t_0_: administration, t_1_: expected peak effect).

To evaluate the short trajectories, we constructed a binary contingency table categorizing medication by effectiveness and HR/RR changes by sign. The chi-square results for HR show no significant association with medication effectiveness (χ^2^(1, n=282) = 1.38, p=0.241). However, the chi-square test was unsuitable for RR data due to the absence of decreasing RRs for ineffective medication, leading to a lack of variability.

Pairwise t-tests (Table 5) revealed significant differences in RR changes following ineffective medication (p=0.017) in a small sample size of n=3. HR trajectories for ineffective interventions approached significance (p=0.068).

**Table 5:** Short-term analysis – Pairwise t-test results for HR and RR changes in response to medication.

|  | HR<br>Sample<br>Size | HR<br>t-value | HR<br>p-value | RR<br>Sample<br>Size | RR<br>t-value | RR<br>p-value |
| --- | --- | --- | --- | --- | --- | --- |
| Effective<br>Medication | 256 | 2.071 | 0.980 | 113 | 3.225 | 0.999 |
| Ineffective<br>Medication | 26 | -1.539 | 0.068 | 3 | -5.302 | 0.017* |

## 4. Discussion

The findings of this study indicate that radar-based VSM can provide an objective measure to support clinical assessment of symptom onset and burden as well as information on the efficacy of treatment plans, particularly through changes in HR and RR. As previously mentioned, an increase in HR/RR is typically observed during periods of distress due to sympathetic activation, followed by a decrease when symptoms are alleviated, reflecting reduced sympathetic activity and enhanced parasympathetic activity[14, 15]. This pattern is reflected in the long-term and short-term analyses conducted in this study.

The long-term analysis revealed that the group mean changes in HR/RR were generally subtle, yet they provided information on vegetative reaction to symptoms and treatment. However, results were slightly more pronounced for symptom exacerbation (HR: +2.8 bpm, RR: +0.7 brpm) compared to amelioration (HR: –1.8 bpm, RR: –0.2 brpm). This difference was supported by significant t-test results for exacerbation (HR: p=0.002, RR: p=0.044), while changes for amelioration were not statistically significant. Chi-square tests suggested a general association between HR and symptom burden (p=0.003), a result reinforced by logistic regression (p=0.001), while RR monitoring showed no significant difference. This may be due to the fact that HR changes, being larger in magnitude than RR changes, were more easily detected by radar, and small measurement errors may have had less impact. Furthermore, RR changes may be less pronounced as seen in radar data since RRs were integrated over longer periods of time and thus dynamic changes may be underrepresented.

GLMM analyses found no significant patient or time dependency. The absence of time dependency is particularly interesting, given the large variations in time intervals between the long-term trajectories. However, as each trajectory was only marked by two timestamps, HR/RR trends might not have been captured effectively, leaving intermediate changes unexamined. Further investigation incorporating three or more time points per trajectory is needed to gain a better understanding of symptom progression.

In the short-term analysis, the group mean HR/RR changes exhibited a similar pattern, with slightly larger changes observed towards an increase of HR and RR, suggesting a sympathetic response to distress when medication was ineffective (HR: +2.0 bpm, RR: +1.0 brpm) compared to a decrease of HR and RR due to a reduction of sympathetic activation as a response to reduced stress, when on-demand medication was effective, leading to symptom amelioration (HR: –0.8 bpm, RR: –0.7 brpm). Pairwise t-tests revealed significant differences in RR changes for ineffective medication (p=0.017), though the small sample size (n=3) limits the statistical power of these results. A larger sample size is required to confirm these findings with greater confidence. Furthermore, our results suggest that HR as a singular vital sign is not a strong marker of medication effectiveness in relieving distress, although a trend (p=0.068) is visible in the ineffective medication group, indicating a potential effect that should be explored further with larger samples or more refined measures. The distribution of HR changes for ineffective medication (see Figure 3), demonstrates that responses were still observed in many cases, with half of the trajectories showing a decline in HR, likely due to the medication’s effects. A further limitation of the study is that the CHR were utilized retrospectively as ground truth for symptom burden and amelioration by treatment. The CHR will reflect on-demand medication, its indication and its effect, but in terms of time stamps, there might be some impreciseness, which may render the radar data analysis less specific and thus lead to an underestimation of effects. Finally, performance of the radar systems utilized in this study was limited in terms of temporal resolution, so that heart rate variability (HRV) as an established biomarker for acute and chronic distress could not be assessed robustly. An increase in radar performance may allow to utilize HRV for increased symptom detection and monitoring.

Overall, our findings suggest that changes in symptom burden are partially reflected in radar-based vital signs, with HR changes showing more prominence than RR changes. In both long-term and short-term analyses, symptom exacerbation or lack of improvement had a more pronounced impact on vital signs than psychometrically proven amelioration. However, factors such as physical activity, which may differ with well-being, and environmental influences likely affected HR and RR, which were not accounted for in this study. A patient may experience symptom relief and then engage in increased activity or interactions, both of which could elevate HR and RR, countering the expected decrease in vital signs. Additionally, the simultaneous occurrence of simultaneous or other symptoms such as nausea or disorientation was not considered.

Despite these challenges, this study demonstrates the high potential of radar-based VSM as a non-invasive, burden-free tool for assessing and monitoring symptoms and symptom burden in PC patients. This technology offers objective support for clinical decision-making alongside patient self-reports and professional assessments by HCPs. However, further research is needed to clarify how medications influence HR and RR changes and how to improve radar performance to record second-tier cardiorespiratory parameters such as HRV and breathing patterns. Larger studies are required to confirm whether radar-based biomarkers reliably reflect medication effectiveness, and to refine radar-based monitoring for better detection of subtle symptom changes.

## Data Availability

The data that support the findings of this study are not openly available due to reasons of privacy and sensitivity and are available from the corresponding author upon reasonable request.

## 6. Authors’ Contribution

T.T.T., and T.S. conceived the study. T.T.T., J.Y., and N.C.A. provided analysis tools. T.T.T., S.G.G., and T.S. analysed the results. T.T.T. created all figures. B.M.E. and A.K. were the technical supervisors, T.S., M.H., and C.O. the clinical supervisors. T.T.T. and T.S. wrote the paper, M.O. substantively revised the paper. All authors commented, critically reviewed, and approved the final paper.

## Acknowledgements

We would like to thank the Palliative Care Department, Universitätsklinikum Erlangen, for providing equipment and assessing the measurements. The research project GUARDIAN was funded by the Deutsche Forschungsgemeinschaft (DFG, German Research Foundation) — SFB 1483 — Project-ID 442419336, EmpkinS.

## References

[1] H. Merlane and L. Cauwood, ‘Core principles of end-of-life care’, Br. J. Nurs., vol. 29, no. 5, pp. 280–282, Mar. 2020, doi: 10.12968/bjon.2020.29.5.280.

[2] L. Radbruch and J. Downing, ‘Principles of palliative care’, Guide Pain Manag. Low-Resour. Settings, vol. 47, 2010.

[3] C. White, D. McMullan, and J. Doyle, ‘“Now that You Mention it, Doctor … “: Symptom Reporting and the Need for Systematic Questioning in a Specialist Palliative Care Unit’, J. Palliat. Med., vol. 12, no. 5, pp. 447–450, May 2009, doi: 10.1089/jpm.2008.0272.

[4] P. Bramati and E. Bruera, ‘Delirium in Palliative Care’, Cancers, vol. 13, no. 23, p. 5893, Nov. 2021, doi: 10.3390/cancers13235893.

[5] S. A. Grossman, V. R. Sheidler, K. Swedeen, J. Mucenski, and S. Piantadosi, ‘Correlation of patient and caregiver ratings of cancer pain’, J. Pain Symptom Manage., vol. 6, no. 2, pp. 53–57, Feb. 1991, doi: 10.1016/0885-3924(91)90518-9.

[6] M. Hodgkins, D. Albert, and L. Daltroy, ‘Comparing patients’ and their physicians’ assessments of pain1’:, Pain, vol. 23, no. 3, pp. 273–277, Nov. 1985, doi: 10.1016/0304-3959(85)90105-8.

[7] C. Pham, K. Poorzarga, M. Nagappa et al., ‘Effectiveness of consumer-grade contactless vital signs monitors: a systematic review and meta-analysis’, J. Clin. Monit. Comput., vol. 36, no. 1, pp. 41–54, Feb. 2022, doi: 10.1007/s10877-021-00734-9.

[8] V. Selvaraju, N. Spicher, J. Wang et al., ‘Continuous monitoring of vital signs using cameras: a systematic review’, Sensors, vol. 22, no. 11, Art. no. 4097, 2022, doi: 10.3390/s22114097.

[9] G. Sun, T. Negishi, T. Kirimoto, T. Matsui, and S. Abe, ‘Noncontact Monitoring of Vital Signs with RGB and Infrared Camera and Its Application to Screening of Potential Infection’, in Non-Invasive Diagnostic Methods – Image Processing, M. Marzec and R. Koprowski, Eds., IntechOpen, 2018. doi: 10.5772/intechopen.80652.

[10] T. P. Naziyok, A. A. Zeleke, and R. Röhrig, ‘Contactless Patient Monitoring for General Wards: A Systematic Technology Review’, Stud. Health Technol. Inform., vol. 228, pp. 707–711, 2016.

[11] M. Liebetruth, K. Kehe, D. Steinritz, and S. Sammito, ‘Systematic Literature Review Regarding Heart Rate and Respiratory Rate Measurement by Means of Radar Technology’, Sensors, vol. 24, no. 3, p. 1003, Feb. 2024, doi: 10.3390/s24031003.

[12] S. G. Grießhammer, A. Malessa, H. Lu et al., ‘Contactless radar-based heart rate estimation in palliative care – a feasibility study and possible use in symptom management’, BMC Palliat. Care, vol. 23, no. 1, p. 273, Nov. 2024, doi: 10.1186/s12904-024-01592-3.

[13] T. Ott, M. Heckel, N. Öhl et al., ‘Palliative care and new technologies. The use of smart sensor technologies and its impact on the Total Care principle’, BMC Palliat. Care, vol. 22, no. 1, p. 50, Apr. 2023, doi: 10.1186/s12904-023-01174-9.

[14] G. Forte, G. Troisi, M. Pazzaglia, V. D. Pascalis, and M. Casagrande, ‘Heart Rate Variability and Pain: A Systematic Review’, Brain Sci., vol. 12, no. 2, p. 153, Jan. 2022, doi: 10.3390/brainsci12020153.

[15] H.-G. Kim, E.-J. Cheon, D.-S. Bai, Y. H. Lee, and B.-H. Koo, ‘Stress and Heart Rate Variability: A Meta-Analysis and Review of the Literature’, Psychiatry Investig., vol. 15, no. 3, pp. 235–245, Mar. 2018, doi: 10.30773/pi.2017.08.17.

[16] Studienregister des Bayerischen Zentrums für Krebsforschung. https://studien.bzkf.de/

[17] S. Stiel, M. E. Matthes, L. Bertram, C. Ostgathe, F. Elsner, and L. Radbruch, ‘Validierung der neuen Fassung des Minimalen Dokumentationssystems (MIDOS2) für Patienten in der Palliativmedizin: Deutsche Version der Edmonton Symptom Assessment Scale (ESAS)’, Schmerz, vol. 24, no. 6, pp. 596–604, Dec. 2010, doi: 10.1007/s00482-010-0972-5.

[18] N. Krumm, S. Stiel, C. Ostgathe et al., ‘Subjektives Befinden bei Palliativpatienten – Ergebnisse der Hospiz-und Palliativerhebung (HOPE)’, Z. Für Palliativmedizin, vol. 9, no. 03, pp. 132–138, Sep. 2008, doi: 10.1055/s-2008-1067515.

[19] F. Michler, K. Shi, S. Schellenberger et al., ‘A Radar-Based Vital Sign Sensing System for In-Bed Monitoring in Clinical Applications’, 2020.

[20] S. Schellenberger, K. Shi, F. Michler, F. Lurz, R. Weigel, and A. Koelpin, ‘Continuous In-Bed Monitoring of Vital Signs Using a Multi Radar Setup for Freely Moving Patients’, Sensors, vol. 20, no. 20, p. 5827, Oct. 2020, doi: 10.3390/s20205827.

[21] S. Linz, F. Lurz, R. Weigel, and A. Koelpin, ‘A review on six-port radar and its calibration techniques’, in 2018 22nd International Microwave and Radar Conference (MIKON), Poznan: IEEE, May 2018, pp. 80–83. doi: 10.23919/MIKON.2018.8405341.

[22] C. Li, V. M. Lubecke, O. Boric-Lubecke, and J. Lin, ‘A Review on Recent Advances in Doppler Radar Sensors for Noncontact Healthcare Monitoring’, IEEE Trans. Microw. Theory Tech., vol. 61, no. 5, pp. 2046–2060, May 2013, doi: 10.1109/TMTT.2013.2256924.

[23] K. Mostov, E. Liptsen, and R. Boutchko, ‘Medical applications of shortwave FM radar: Remote monitoring of cardiac and respiratory motion’, Med. Phys., vol. 37, no. 3, pp. 1332–1338, Mar. 2010, doi: 10.1118/1.3267038.

[24] C. Will, K. Shi, S. Schellenberger et al., ‘Local Pulse Wave Detection Using Continuous Wave Radar Systems’, IEEE J. Electromagn. RF Microw. Med. Biol., vol. 1, no. 2, pp. 81–89, Dec. 2017, doi: 10.1109/JERM.2017.2766567.

[25] C. Will, K. Shi, S. Schellenberger et al., ‘Radar-Based Heart Sound Detection’, Sci. Rep., vol. 8, no. 1, p. 11551, Jul. 2018, doi: 10.1038/s41598-018-29984-5.

[26] D. Springer, L. Tarassenko, and G. Clifford, ‘Logistic Regression-HSMM-based Heart Sound Segmentation’, IEEE Trans. Biomed. Eng., pp. 1–1, 2015, doi: 10.1109/TBME.2015.2475278.

[27] K. Shi, S. Schellenberger, C. Will et al., ‘A dataset of radar-recorded heart sounds and vital signs including synchronised reference sensor signals’, Sci. Data, vol. 7, no. 1, p. 50, Feb. 2020, doi: 10.1038/s41597-020-0390-1.

[28] L. Herzer, A. Muecke, R. Richer et al., ‘Influence of Sensor Position and Body Movements on Radar-Based Heart Rate Monitoring’, in 2022 IEEE-EMBS International Conference on Biomedical and Health Informatics (BHI), Ioannina, Greece: IEEE, Sep. 2022, pp. 1–4. doi: 10.1109/BHI56158.2022.9926775.

[29] V. Chandola, A. Banerjee, and V. Kumar, ‘Anomaly detection: A survey’, ACM Comput. Surv., vol. 41, no. 3, pp. 1–58, Jul. 2009, doi: 10.1145/1541880.1541882.

[30] P. J. Rousseeuw and C. Croux, ‘Alternatives to the Median Absolute Deviation’, J. Am. Stat. Assoc., vol. 88, no. 424, pp. 1273–1283, Dec. 1993, doi: 10.1080/01621459.1993.10476408.

[31] B. Iglewicz and D. C. Hoaglin, How to detect and handle outliers. in The ASQC basic references in quality control: statistical techniques, no. 16. Milwaukee, Wis: ASQC Quality Press, 1993.

